# Impact of non-pharmaceutical interventions for SARS-CoV-2 on norovirus outbreaks: an analysis of outbreaks reported by 9 US States

**DOI:** 10.1101/2020.11.25.20237115

**Authors:** Alicia N.M. Kraay, Peichun Han, Anita K. Kambhampati, Mary E. Wikswo, Sara A. Mirza, Benjamin A. Lopman

## Abstract

**Importance:** The impact of non-pharmaceutical interventions (NPIs) in response to the SARS-CoV-2 pandemic on incidence of other infectious diseases is still being assessed.

**Objective:** To determine if the observed change in reported norovirus outbreaks in the United States was best explained by underreporting, seasonal trends, or reduced exposure due to NPIs. We also aimed to assess if the change in reported norovirus outbreaks varied by setting.

**Design:** An ecologic, interrupted time series analysis of norovirus outbreaks from nine states reported to the National Outbreak Reporting System (NORS) from July 2012–July 2020.

**Setting:** Surveillance data from Massachusetts, Michigan, Minnesota, Ohio, Oregon, South Carolina, Tennessee, Virginia, and Wisconsin were included in the analysis.

**Participants:** 9,226 reports of acute gastroenteritis outbreaks with norovirus as an epidemiologically suspected or laboratory-confirmed etiology were included in the analysis, resulting in more than 8 years of follow up. Outbreak reports from states that participated in NoroSTAT for at least 4 years were included in the analysis (range: 4–8 years).

**Exposure:** The main exposure of interest was time period: before (July 2012–February 2020) or after (April 2020–July 2020) the start of NPIs in the United States

**Main outcome:** The main outcome of interest was monthly rate of reported norovirus outbreaks. As a secondary outcome, we also examined the average outbreak size.

**Results:** We found that the decline in norovirus outbreak reports was significant for all 9 states considered (pooled incidence rate ratio (IRR) comparing April 2020-July 2020 vs. all pre-COVID months for each state= 0.14, 95% CI: 0.098, 0.21; *P*=<0.0001), even after accounting for typical seasonal decline in incidence during the summer months. These patterns were similar across a variety of settings, including nursing homes, child daycares, healthcare settings, and schools. The average outbreak size was also reduced by 61% (95% CI: 56%, 42.7%; *P*=<0.0001), suggesting that the decline does not reflect a tendency to report only more severe outbreaks due to strained surveillance systems, but instead reflects a decline in incidence.

**Conclusions and relevance:** While NPIs implemented during the spring and summer of 2020 were intended to reduce transmission of SARS-CoV-2, these changes also appear to have impacted the incidence of norovirus, a non-respiratory pathogen. These results suggest that NPIs may provide benefit for preventing transmission of other human pathogens, reducing strain to health systems during the continued SARS-CoV-2 pandemic.

**Disclaimer:** The findings and conclusions in this report are those of the authors and do not necessarily represent the official position of the US Centers for Disease Control and Prevention (CDC).

---

Since March 2020, non-pharmaceutical interventions (NPIs) such as social distancing, mask wearing, surface disinfection, and increased hand hygiene have become widespread throughout the United States and worldwide in an effort to reduce transmission of severe acute respiratory syndrome coronavirus 2 (SARS-CoV-2)^1^. These changes likely reduced the incidence of influenza in other countries ^2,3^, and may also influence the transmission of other pathogens like norovirus. At the same time, healthcare-seeking declined dramatically for acute illnesses, including some illnesses for which incidence would not have been expected to be linked to NPIs ^4^, suggesting that some decrease in incidence, even for life-threatening diseases, could be related to underreporting. NPIs for SARS-CoV-2 prevention began near the end of the main transmission season for winter-time pathogens, including norovirus ^5^, so careful analysis is required to separate the effect of responses directed at SARS-CoV-2 from seasonal patterns.

Starting in spring of 2020, there was a large drop in the number of norovirus outbreaks reported to the Centers for Disease Control and Prevention (CDC). We used Poisson regression models to investigate if this decrease was related to NPIs or could be explained by seasonal changes in incidence and/or underreporting.

## Methods

State and local health departments report outbreaks of enteric illness to CDC using the National Outbreak Reporting System (NORS). In August 2012, the Norovirus Sentinel Testing and Tracking network (NoroSTAT) was established to improve timeliness and completeness of norovirus outbreak reporting to CDC. NoroSTAT states are required to report all epidemiologically suspected and laboratory-confirmed norovirus outbreaks within 7 business days of identification and provide minimum outbreak data elements including the setting and number of cases for each outbreak.^6,7^ NoroSTAT began with 5 states and expanded to 12 states by 2019 (**Table 1**). Participating states were chosen from those with the highest per capita norovirus outbreak reporting rates and were less likely to be affected by underreporting. Three states participated in NoroSTAT for less than 3 surveillance years prior to 2020 and were therefore not included in our analysis. We used data from the remaining 9 states, beginning the first reporting year of each state’s participation in NoroSTAT and concluding in July 2020 (the end of the 2020 reporting year).

**Table 1.** Average monthly norovirus outbreaks before and during the SARS-CoV-2 pandemic. While all months were included in the full GAM analysis (in Figure 1), only the months April–July are shown in this table to illustrate descriptive trends for similar times of year, without formally adjusting for seasonality.

| Participating state | Date state joined NoroSTAT | Pre-SARS-CoV-2 |  | SARS-CoV-2 | Monthly norovirus outbreaks April–July 2020 vs. monthly outbreaks reference period<br>Crude IRR (95% CI) |
| --- | --- | --- | --- | --- | --- |
|  |  | Reference Period (April 1–July 31 of each year) | Monthly norovirus outbreaks during the reference period<br>Average (Range) | Monthly norovirus outbreaks April–July 2020<br>Average (Range) |  |
| Minnesota | August 1, 2012 | 2013–2019 | 5.82 (0, 16) | 0.75 (0, 2) | 0.129 (0.041, 0.404) |
| Ohio | August 1, 2012 | 2013–2019 | 8.36 (1, 31) | 0.50 (0, 2) | 0.060 (0.015, 0.241) |
| Oregon | August 1, 2012 | 2013–2019 | 7.93 (2, 22) | 1.25 (0, 3) | 0.158 (0.065, 0.383) |
| Tennessee | August 1, 2012 | 2013–2019 | 2.56 (0, 10) | 0 (0, 0) | 0 (0, Inf) |
| Wisconsin | August 1, 2012 | 2013–2019 | 8.96 (2, 34) | 2 (1, 4) | 0.223 (0.110, 0.451) |
| Michigan | August 1, 2015 | 2016–2019 | 6.88 (0, 20) | 0.50 (0, 1) | 0.073 (0.018, 0.294) |
| South Carolina | August 1, 2015 | 2016–2019 | 3.75 (1, 10) | 0.50 (0, 2) | 0.133 (0.033, 0.545) |
| Massachusetts | August 1, 2016 | 2017–2019 | 5.67 (1, 18) | 0.75 (0, 3) | 0.132 (0.042, 0.421) |
| Virginia | August 1, 2016 | 2017–2019 | 5.75 (0, 21) | 0.75 (0, 2) | 0.130 (0.041, 0.414) |
| Average | -- | -- | 6.19 (0, 34) | 0.78 (0, 4) | 0.126 (0.087, 0.18) |

To assess how NPIs implemented in response to SARS-CoV-2 might have impacted norovirus outbreak incidence, we compared the number of monthly outbreaks before (August 2012–February 2020) and after NPIs began (April–July 2020), adjusting for seasonal changes in incidence using cyclic cubic generalized additive model (GAM) smoothing terms.^8^ We also compared the difference in reported outbreaks by month to assess monthly variation in incidence after NPIs began. March 2020 was excluded from the analysis because it was a transition month, during which social distancing measures (e.g., stay at home orders, school and restaurant closures) and recommendations for improved infection prevention strategies (including increased hand hygiene and surface disinfection) were implemented throughout the United States. The same pre/post period was used for all states because reductions in mobility, a proxy for social distancing intensity,^9^ started at about the same time, reaching a maximum reduction by April (see **Appendix Figure S1**). By April 2020, all 9 of the included states had implemented their initial NPIs. Because norovirus seasonality varied by both state and setting, we ran separate models (1) for each state (combining all settings) and (2) for each setting (combining all states). Setting-specific models were adjusted for state using indicator variables and adjusted for season intrinsically by using only data from April—July of each year.

For state-specific models, we considered two alternative ways of adjusting for seasonality in the pooled effect estimate across states: (1) we combined all states into a single meta-regression model, where seasonally-adjusted state-specific estimates were weighted by inverse variance with a random effect for state and (2) we subset the data to only include April–July of each year just like the setting-specific models (since the reporting year begins in August, this gave us one fewer year of surveillance data for each state) and used standard Poisson models to estimate the impact of NPIs. The meta-regression approach (our primary analysis) allows the timing and degree of seasonality to vary by state, which should minimize residual confounding by season due to state-level differences.

To test whether the decline in norovirus outbreaks was due to changes in reporting, particularly preferential reporting of only the largest outbreaks, we compared the size of outbreaks reported to NoroSTAT during the spring and summer months prior to the start of the SARS-CoV-2 pandemic (April–July 2013–2019) with outbreak size after the pandemic began (April–July 2020) using Poisson regression models. Because these models were stratified to compare the same times of year, they were not adjusted for month. We also assessed whether changes in outbreak size varied by setting, comparing outbreaks in nursing homes, child daycares, schools/colleges/universities, healthcare facilities, and other locations (see **Appendix Table S1** for detailed setting information).

All code used in the analysis is available on github. All statistical analysis was conducted in R version 4.0.2. We used the ‘mgcv’ R package for GAM regression models and the ‘metafor’ package to obtain the pooled estimate across states.

## Results

Across all states, there was a monthly mean and range of 6.19 (0 to 34) reported outbreaks in April–July during the reference period for each state (range: 3-7 years) compared to 0.78 (0 to 4) during April–July 2020 (**Table 1**). In the full GAM model, accounting for seasonal trends, the incidence of reported norovirus outbreaks was significantly reduced for all states April–July 2020 compared with the reference period for each state (pooled monthly incidence rate ratio (IRR)=0.14, 95% CI: 0.098, 0.21) (**Figure 1A**). The point estimate was similar when states were combined, without using meta-regression (IRR=0.121, 95% CI: 0.083, 0.18) and was also similar for the model that included only April–July of each year (IRR=0.126, 95% CI: 0.087, 0.18). Point estimates for all states were strong and effect sizes were similar, with IRR values ranging from 0 (95% CI: 0, Inf) for Tennessee to 0.223 (95% CI: 0.11, 0.45) for Wisconsin. Likewise, the reduction in outbreak incidence was apparent in all settings but was least dramatic in long-term care and other healthcare facilities (**Figure 1B**). In general, the decline in monthly outbreaks began in April, after NPIs had been more fully implemented. Beginning in April, the number of reported monthly outbreaks dropped to nearly zero for all states and remained at low levels through the end of July (**Figure 2**). In terms of size, outbreaks were on average 61% smaller in April–July 2020 (95% CI: 56%, 66%) compared with the same period in prior years, with similar patterns across all settings (**Figure 3**). The exception was in settings classified as “other”, for which the single outbreak that occurred between April and July was similar to the average outbreak size in prior years.

**Figure 1.**
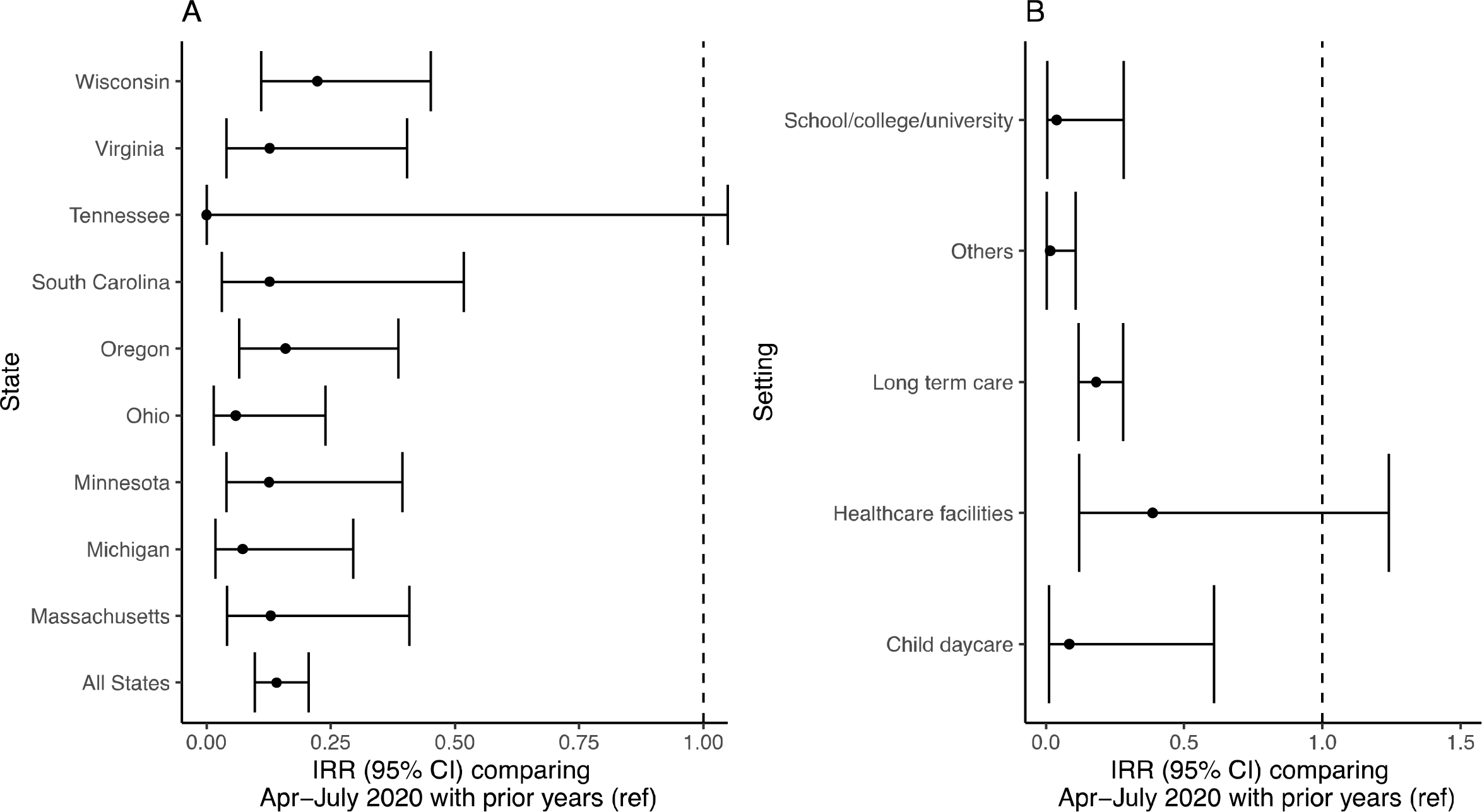
Incidence rate ratio and 95% confidence intervals of norovirus outbreaks for April– July 2020 compared with prior years (ref) by A) state and B) outbreak setting. Note that there were no outbreaks reported in Tennessee in April–July 2020.

**Figure 2.**
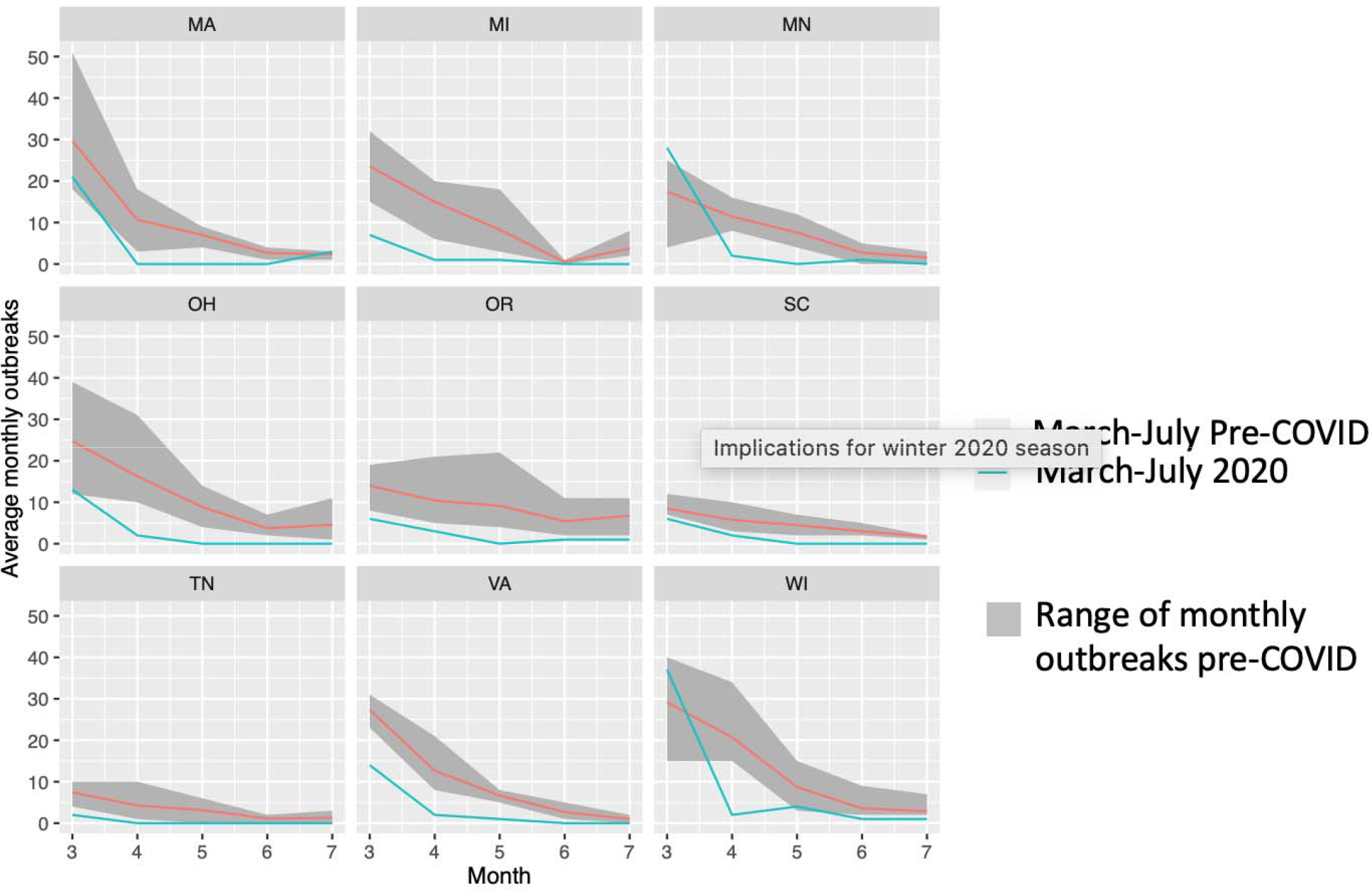
Average number of norovirus outbreaks in March–July during the reference period (red) and March–July 2020 (blue). The grey ribbon shows the range of monthly outbreaks observed in the reference period for each state.

**Figure 3.**
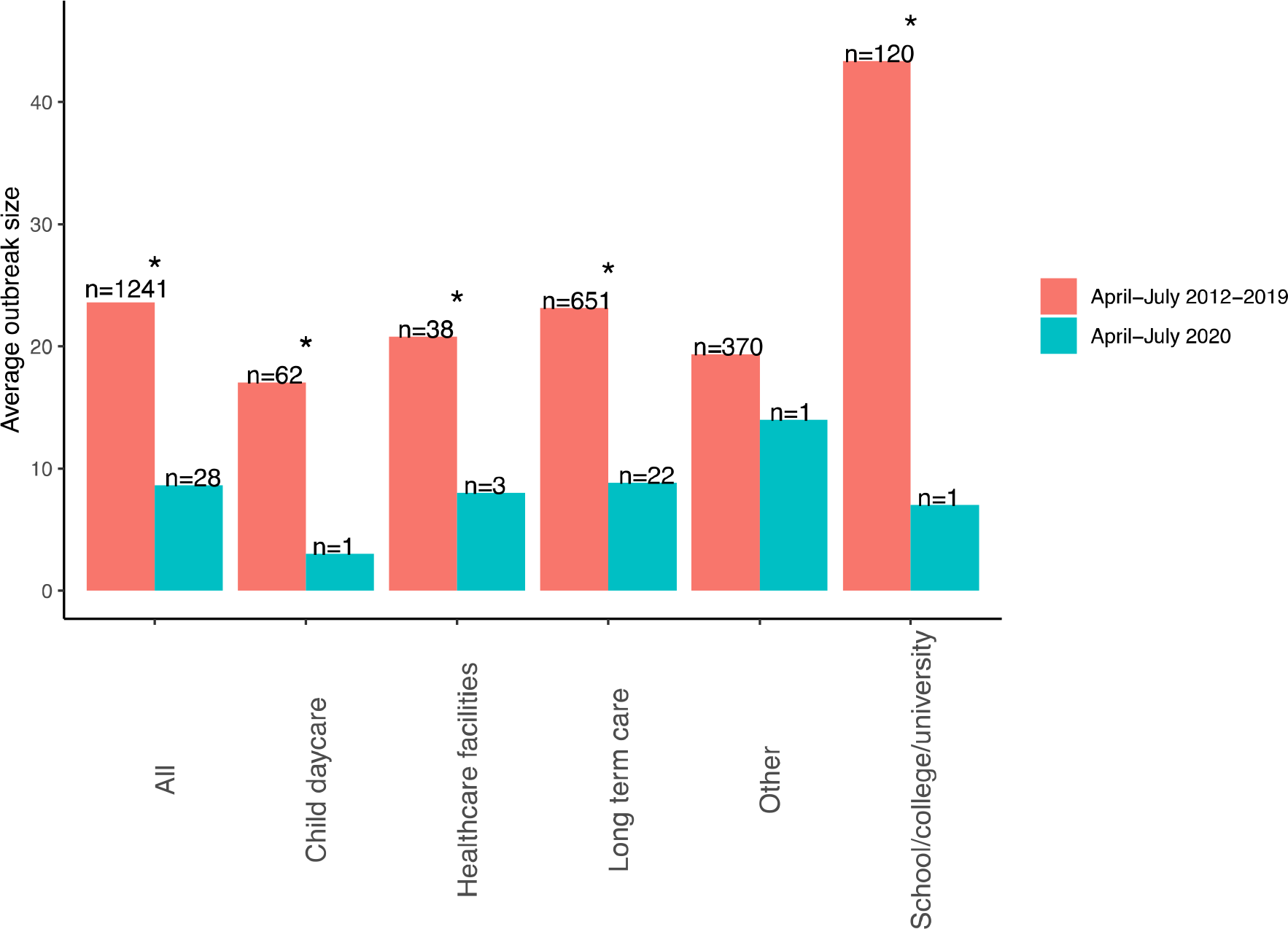
Average norovirus outbreak size by setting for April–July 2013-2019 (red) and for April–July 2020 (blue). An asterisk (*) indicates a significant difference between time periods using Poisson regression. The number of outbreaks (n) used to calculate the size for each period is shown above each colored bar.

## Discussion

We found a decline of >80% in incidence of reported norovirus outbreaks during April– July 2020 compared to the same time period in previous years, likely in part due to the implementation of NPIs for COVID-19 in spring and summer 2020. This decline was consistent across states and settings and could not be explained solely by seasonality. Given that this decline coincided with a decrease in average outbreak size, underreporting is also unlikely to be the primary explanation for these findings. Moreover, based on ongoing dialogue with state public health departments participating in NoroSTAT, all known norovirus outbreaks continue to be reported to NORS.

While smaller and fewer outbreaks were reported across all settings, the difference was least dramatic in nursing homes and healthcare facilities. Stringent SARS-CoV-2 prevention measures have been implemented in nursing homes and healthcare facilities throughout the United States in response to the COVID-19 pandemic^10,11^, yet unlike schools and daycares, these facilities could not completely close.

While uncertain, differences in effect sizes suggest that these patterns may initially have been driven by widespread closures of locations like schools where norovirus outbreaks commonly occur. As the 2020-2021 norovirus season begins and locations such as schools and restaurants continue to reopen, ongoing patterns of norovirus incidence based on outbreak surveillance might provide more clarity on the reasons behind these trends. For example, as schools, daycares, and restaurants begin to reopen, incidence may increase again^12^. However, sustained social distancing, more frequent surface disinfection, and enhanced hand hygiene might continue to reduce the number of norovirus outbreaks. Conversely, population immunity may be lower at the start of the norovirus season, which could lead more frequent or larger norovirus outbreaks.

Our findings are similar to reported declines in norovirus incidence in the England and Wales during March and April 2020 ^13^. While most initial studies showing declining infectious disease trends have focused on influenza, we show that these declines in incidence were observed for norovirus, a non-respiratory pathogen. Unlike SARS-CoV-2 and other respiratory pathogens, which are commonly spread by droplets^14^, norovirus is primarily transmitted by the fecal-oral route^15^. Therefore, the unanticipated benefits of NPIs for SARS-CoV-2 might also be present for other pathogens transmitted person-to-person.

## Data Availability

Norostat data are publicly available at https://www.cdc.gov/norovirus/reporting/norostat/data-table.html. More detailed Norostat data can be obtained by sending a formal data request to the Centers for Disease Control and Prevention at.

