## Appendix for "Impact of non-pharmaceutical interventions for SARS-CoV-2 on norovirus outbreaks: an analysis of outbreaks reported by 9 US States"

**Figure S1.** Mobility relative to the median value for trips taken from January 3–February 6, 2020 for trips to A) workplaces or B) retail/recreation for the nine NoroSTAT states included in the analysis. Different states are shown in different line colors. Data are publicly available at: <https://github.com/ActiveConclusion/COVID19_mobility/tree/master/google_reports>

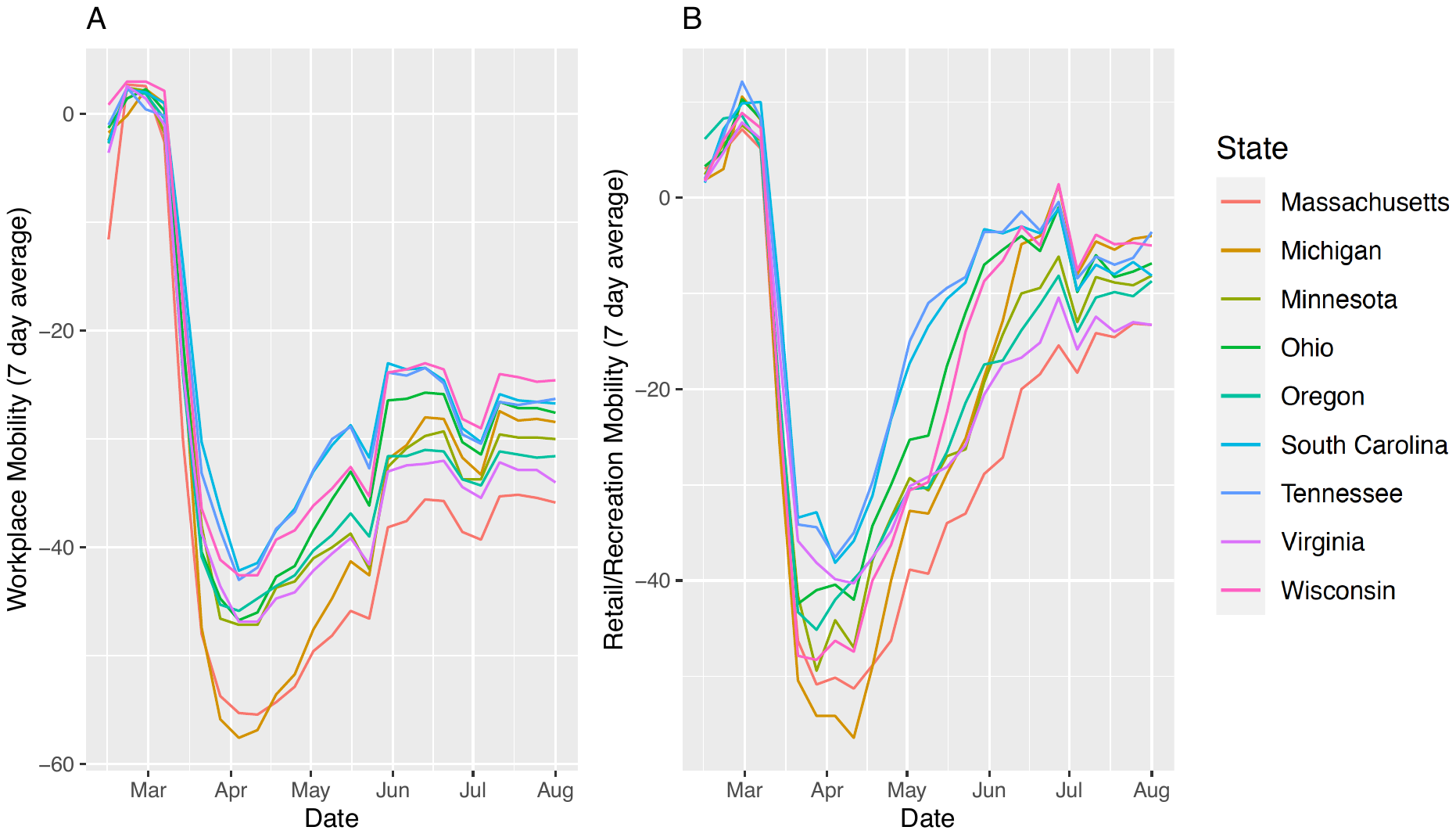

**Table S1.** More detailed description of outbreaks in ‘other’ locations

| Location | August 2012–February 2020  N=370  % (number of outbreaks) | April–July 2020  N=1  % (number of outbreaks) |
| --- | --- | --- |
| Restaurant | 38% (139) | 0% (0) |
| Other (no details provided) | 13% (47) | 0% (0) |
| Camps/cabins | 11% (40) | 0% (0) |
| Private residence | 10% (37) | 0% (0) |
| Banquet facilities/caterers/other food | 6% (24) | 0% (0) |
| Office/indoor workplace | 5% (19) | 0% (0) |
| Multiple (no details provided) | 5% (17) | 0% (0) |
| Prison | 3% (12) | 0% (0) |
| Event space | 2% (8) | 0% (0) |
| Hotel/Motel | 2% (7) | 0% (0) |
| Religious facility | 2% (6) | 100% (1) |
| Unknown | 1% (5) | 0% (0) |
| Festival/fair | 1% (4) | 0% (0) |
| Park | 1% (3) | 0% (0) |
| Shelter/group home/transitional housing | 1% (2) | 0% (0) |
